# Famotidine Use is Associated with Improved Clinical Outcomes in Hospitalized COVID-19 Patients: A Propensity Score Matched Retrospective Cohort Study

**DOI:** 10.1101/2020.05.01.20086694

**Authors:** Daniel E. Freedberg, Joseph Conigliaro, Magdalena E. Sobieszczyk, David D. Markowitz, Aakriti Gupta, Max R. O’Donnell, Jianhua Li, David A. Tuveson, Zhezhen Jin, William C. Turner, Donald W. Landry, Timothy C. Wang, Kevin J. Tracey, Michael V. Callahan, Julian A. Abrams

## Abstract

**Background and Aims:** No medications are proven to improve clinical outcomes in COVID-19. Famotidine is commonly used for gastric acid suppression but has recently gained attention as an antiviral that may inhibit SARS-CoV-2 replication. This study tested whether famotidine use is associated with improved clinical outcomes in patients with COVID-19 initially hospitalized to a non-intensive care setting.

**Methods:** This was retrospective cohort study conducted among consecutive hospitalized patients with COVID-19 infection from February 25 to April 13, 2020 at a single medical center. The primary exposure was famotidine, received within 24 hours of hospital admission. The primary outcome was intubation or death. Propensity score matching was used to balance the baseline characteristics of patients who did and did not use famotidine.

**Results:** 1,620 hospitalized patients with COVID-19 were identified including 84 (5.1%) who received famotidine within 24 hours of hospital admission. 340 (21%) patients met the study composite outcome of death or intubation. Use of famotidine was associated with reduced risk for death or intubation (adjusted hazard ratio (aHR) 0.42, 95% CI 0.21-0.85) and also with reduced risk for death alone (aHR 0.30, 95% CI 0.11-0.80). After balancing baseline patient characteristics using propensity score matching, these relationships were unchanged (HR for famotidine and death or intubation 0.43, 95% CI 0.21-0.88). Proton pump inhibitors, which also suppress gastric acid, were not associated with reduced risk for death or intubation.

**Conclusion:** Famotidine use is associated with reduced risk of intubation or death in hospitalized COVID-19 patients. Randomized controlled trials are warranted to determine whether famotidine therapy improves outcomes in hospitalized COVID-19 patients.

## INTRODUCTION

COVID-19 caused 2 million cases and over 150,000 deaths worldwide as of mid-April 2020 (1). Clinical trials are underway to assess the efficacy of a variety of antiviral drugs. However, many of these drugs have toxicities and thus far no drug has been proven to improve outcomes in COVID-19 patients.

Famotidine is a histamine-2 receptor antagonist that suppresses gastric acid production. *In vitro*, famotidine inhibits HIV replication (2). Recently, Wu *et al*. (3) used computational methods to predict structures of proteins encoded by the SARS-CoV-2 genome and identified famotidine as one of the drugs most likely to inhibit the 3-chymotrypsin-like protease (3CL^pro^) which processes proteins essential for viral replication (4). We hypothesized that famotidine would be associated with improved clinical outcomes among hospitalized patients with COVID-19. To explore this, we performed a retrospective cohort study at a single academic center located at the epicenter of the COVID-19 pandemic in the United States.

## METHODS

### Population

Adults aged 18 years or more were eligible for the study if they were admitted to Columbia University Irving Medical Center or its affiliate the Allen Pavilion from February 25, 2020 to April 13, 2020 and tested positive for SARS-CoV-2 by nasopharyngeal polymerase chain reaction at presentation or within no more than 72 hours following admission. This 72-hour window was selected because, during the earliest phase of the SARS-CoV-2 pandemic, testing availability was limited and could take up to 72 hours for a result. Patients were excluded if they survived less than 48 hours following hospital admission or if they required urgent or semi-urgent intubation within 48 hours of hospital admission. This study was approved by the institutional review board of the Columbia University Irving Medical Center.

### Exposure

The primary exposure was use of famotidine, classified as present if famotidine was received within 24 hours of hospital admission and otherwise classified as absent. Famotidine use was ascertained directly from electronic medical order entry records and could be intravenous or oral, at any dose or duration. Home use of famotidine was examined to understand the reason underlying in-hospital use of famotidine and was classified based on electronic medication reconciliation performed at the time of hospital admission.

### Primary outcome

The primary outcome was a composite of death or endotracheal intubation within 30 days of hospital admission (intubation-free survival). Mortality data was ascertained from the electronic medical record (EMR), which interfaces with the social security death index. Endotracheal intubation was ascertained from EMR documentation of need for mechanical ventilation. The rationale for the combined primary outcome was twofold: 1) many patients who deteriorated clinically died without being intubated, often due to transition to palliative care; 2) hospitalization stays for intubated COVID-19 patients have been very long, and many intubated COVID-19 patients at the time of the analyses may ultimately not survive.

### Co-variables

Based on emerging reports of risk factors for COVID-19, the following co-variables were selected for inclusion in the analysis: pre-existing diabetes, hypertension, coronary artery disease (CAD), heart failure, end-stage renal disease or chronic kidney disease, and chronic pulmonary disorders, all classified based on the presence of corresponding ICD-10 codes at the time of hospital admission; obesity, classified based on body mass index (BMI); and age, classified as <50 years old, 50-65 years old, and >65 years old. To assess severity of COVID-19, the first recorded form of supplemental oxygen after triage was captured and was classified as room air, nasal cannula oxygen, or non-rebreather/similar. Use of proton pump inhibitors was classified in the same manner as use of famotidine so that proton pump inhibitors could be evaluated to test whether any effects of famotidine might be related to acid suppression. The maximum value of plasma ferritin was obtained during the study period for each patient to use as a surrogate for the extent of cytokine storm (normal laboratory range 13.0 to 150.0 ng/mL).

### Statistical approach

Categorical variables were compared across exposure groups using chi-squared tests. Full and reduced Cox proportional hazards models were constructed within the complete cohort, with patients followed from the time of hospital admission until the first of the following events: death, intubation, 30 days of follow-up, or the close of the study on April 20, 2020. Because patients were excluded if they died or were intubated prior to hospital day 2, this effectively meant that patients were followed from day 2 to day 30. This design was selected to avoid immortal time bias (i.e., because the exposure was classified based on the 24 hour period after hospitalization and the at-risk period did not begin until hospital day 2). Cox proportional hazards modeling was performed on the full cohort, and a matched subset was examined with propensity scoring matching to balance baseline characteristics based on use of famotidine. This provided the opportunity for a minimum of 7 days of follow-up time for all patients in the study. The proportional hazards assumption was verified by visual inspection of time-to-event data and by testing for a non-zero slope in the Schoenfeld residuals (5). The full Cox model included all baseline variables. For the reduced model, variables were dropped stepwise unless they had a significant independent relationship with the composite outcome or unless they altered the β-coefficient representing famotidine by at least 10%. Propensity score matching was then performed to balance the baseline characteristics of patients with respect to use of famotidine with a 5:1 nearest-neighbor matching strategy and a caliper of 0.2. The primary analysis was conducted as a time-to-event model within the propensity score-matched cohort, using the same approach. All analyses were performed using STATA statistical software (version 14, StataCorp) at the *α* = .05 level of significance.

### Additional analyses

Several sensitivity analyses were performed. First, use of proton pump inhibitors was compared to no proton pump inhibitors within the complete (unmatched) cohort after excluding those who used famotidine. The purpose of this analysis was to test whether unmeasured patient characteristics related to use of acid suppression rather than famotidine were associated with improved outcomes in COVID-19. Second, an additional study cohort was built including records from patients who tested negative for SARS-CoV-2 during the study period. Within this cohort, use of famotidine was compared to no famotidine to test whether unmeasured patient characteristics related to use of famotidine were associated with improved outcomes regardless of reason for hospitalization (i.e., to test whether the observed association with famotidine was specific for COVID-19 patients).

## RESULTS

### Population and use of famotidine

1,620 patients met criteria for analysis including 84 patients (5.1%) who received famotidine within 24 hours of hospital admission. Home use of famotidine was documented on admission medication reconciliation in 15% of those who used famotidine while hospitalized compared to 1% of those who did not (p<0.01). 28% of all famotidine doses were intravenous; 47% were 20 mg, 35% were 40 mg, and 17% were 10 mg. Famotidine users received a median 5.8 days of drug for a total median dose of 136 mg (63 – 233 mg). There were minimal differences comparing patients who used famotidine to those who did not, and balance between the groups was further improved after propensity score matching (**Table 1**).

### Death or intubation

142 (8.8%) patients were intubated and 238 (15%) died; 340 (21%) patients met the composite study outcome. In crude analysis, use of famotidine was significantly associated with reduced risk for the composite outcome of death or intubation (**Figure 1A**, log-rank p<0.01). This association was driven primarily by the relationship between famotidine and death (**Figure 1B**, log-rank p<0.01) and when those who died prior to intubation were excluded, there was no association between use of famotidine and intubation (log-rank p=0.40). After adjusting for baseline patient characteristics, use of famotidine remained independently associated with risk for death or intubation (**Table 2**, adjusted hazard ratio (aHR) 0.42, 95% CI 0.21-0.85) and this remained unchanged after propensity score matching to further balance the co-variables (HR 0. 43, 95% CI 0.21-0.88).

**Figure 1.**
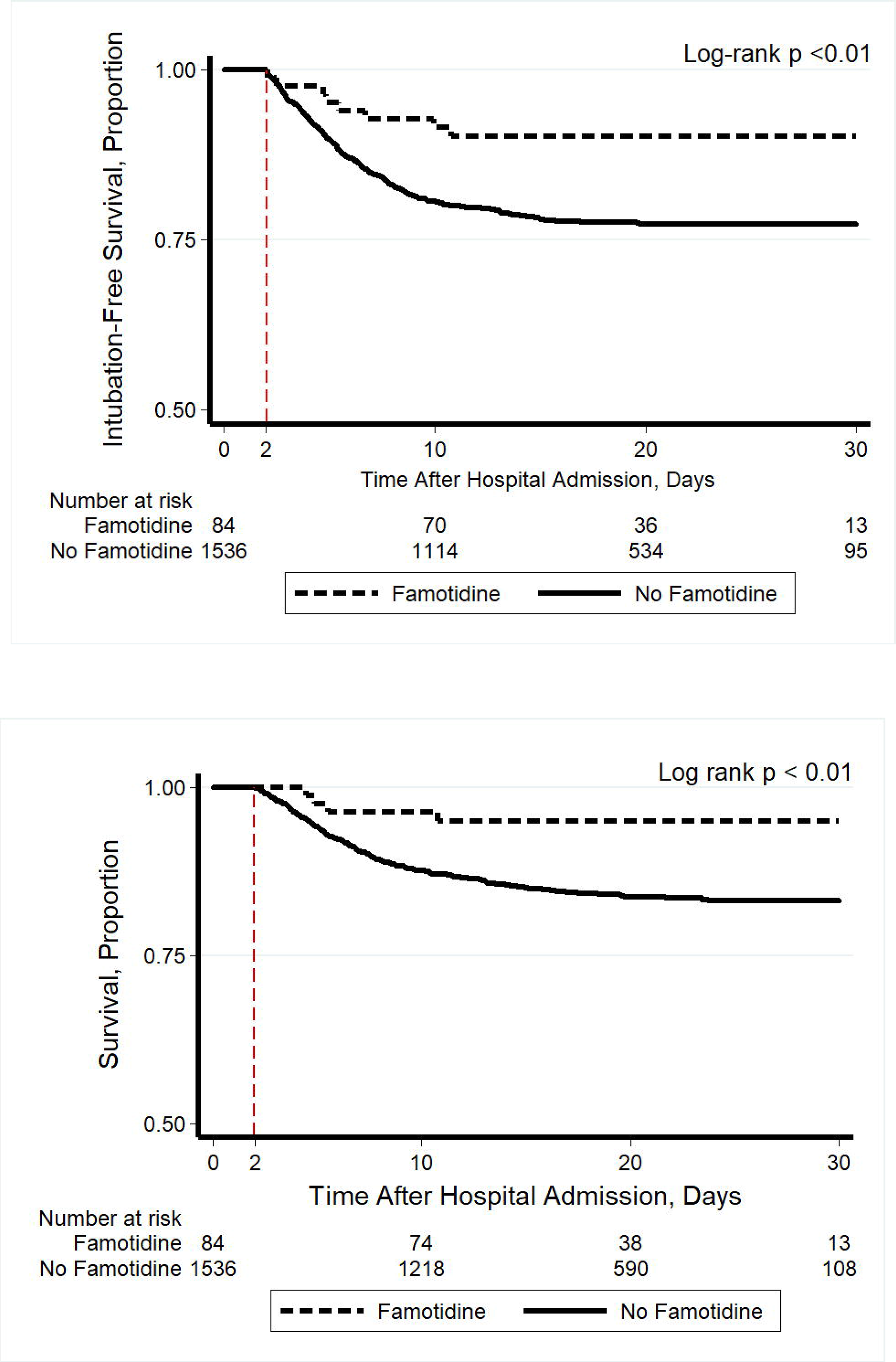
Kaplan-Meier plot showing (A) intubation-free survival and (B) survival through a maximum of 30 days after hospital admission, stratified by use of famotidine. Patients were included in the study if they survived without intubation for two days following hospital admission. Use of famotidine was classified as present if it was received within the first 24 hours following hospital admission (any dose, form of administration, or duration) and otherwise as absent. The at-risk time began on hospital day 2 (indicated with a dashed red line) and patients were followed until hospital day 30. This study design avoided potential for immortal time bias because the exposure was classified prior to the start of the at-risk period.

### Additional analyses

Use of proton pump inhibitors (PPIs) was analyzed because PPIs are also gastric acid suppression medications with similar indications as famotidine. There was a no protective effect associated with use of PPIs (aHR 1.34, 95% CI 1.06-1.69). Next, 784 patients without COVID-19 who were hospitalized during the same study period were analyzed; among these patients, famotidine was not associated with reduced risk for death or intubation (24 deaths or intubations, log-rank p=0.70). The maximum plasma ferritin value during the hospitalization was assessed to address the hypothesis that, by blocking viral replication, famotidine reduces cytokine storm during COVID-19. Median ferritin was 708 ng/mL (IQR 370-1,152) among users of famotidine versus 846 ng/mL (IQR 406-1,552) among non-users (rank-sum p=0.03).

## CONCLUSIONS

This retrospective study found that, in patients hospitalized with COVID-19, famotidine use was associated with a reduced risk of clinical deterioration leading to intubation or death. The study was premised on the assumption that use of famotidine represented a continuation of home use, but documentation of why famotidine was given was poor. The results were specific for famotidine (no protective association was seen for PPIs) and also specific for COVID-19 (no protective association in patients without COVID-19). A lower peak ferritin value was observed among users of famotidine, supporting the hypothesis that use of famotidine may decrease cytokine release in the setting of SARS-CoV-2 infection. A randomized controlled trial is currently underway to determine whether famotidine can improve clinical outcomes in hospitalized COVID-19 patients (NCT04370262).

Famotidine has not previously been studied in patients for antiviral effects, and there is limited relevant prior data. An untargeted computer modeling analysis identified famotidine as one of the highest-ranked matches for drugs predicted to bind 3CL^pro^ (3), a SARS-CoV-2 protease which generates non-structure proteins critical to viral replication (4). In the 1990s, histamine-2 receptor antagonists including famotidine were shown to inhibit HIV replication without affecting lymphocyte viability *in vitro* (2, 6, 7).

There are limitations to the study. It was observational, and we cannot exclude the possibility of unmeasured confounders or hidden bias that account for the association between famotidine use and improved outcomes. No samples were gathered, and mechanism cannot be directly assessed. Finally, this was a single center study, which may limit generalizability of the findings.

In sum, in patients hospitalized with COVID-19 and not initially intubated, famotidine use was associated with a two-fold reduction in clinical deterioration leading to intubation or death. These findings are observational and should not be interpreted to mean that famotidine has a protective effect against COVID-19. Randomized controlled trials are underway.

## Data Availability

Data will be made available by the corresponding author upon request, with appropriate safeguards for patient privacy.

## Acknowledgments

The authors thank Dr. Michael Wigler and Dr. Richard Axel for useful suggestions.

---

**Supplemental Table 1.** Patient characteristics at the time of hospital admission for COVID-19, stratified by use of famotidine.

| Characteristics | Complete Cohort |  |  | After Propensity Score Matching |  |  |
| --- | --- | --- | --- | --- | --- | --- |
|  | Famotidine<br>(n=84) | No Famotidine<br>(n=1,536) | p-value | Famotidine<br>(n=84) | No Famotidine<br>(n=420) | p-value |
| Age |  |  | 0.39 |  |  | 0.51 |
| <50 years old | 13 (15%) | 320 (21%) |  | 13 (15%) | 57 (14%) |  |
| 50-65 years old | 31 (37%) | 483 (31%) |  | 31 (37%) | 184 (44%) |  |
| >65 years old | 40 (48%) | 733 (48%) |  | 40 (48%) | 179 (43%) |  |
| Female sex | 45 (54%) | 864 (56%) | 0.63 | 39 (46%) | 208 (50%) | 0.60 |
| Race/ethnicity |  |  | 0.20 |  |  | 0.90 |
| Hispanic | 25 (30%) | 601 (39%) |  | 25 (30%) | 127 (30%) |  |
| White, non-hispanic | 19 (23%) | 336 (22%) |  | 19 (23%) | 82 (20%) |  |
| Black, non-hispanic | 18 (21%) | 322 (21%) |  | 18 (21%) | 102 (24%) |  |
| Other | 22 (26%) | 277 (18%) |  | 22 (26%) | 109 (26%) |  |
| BMI, kg/m<sup>2</sup> |  |  | 0.17 |  |  | 0.97 |
| <25.0 | 15 (18%) | 295 (19%) |  | 15 (18%) | 66 (16%) |  |
| 25.0-29.9 (overweight) | 30 (36%) | 388 (25%) |  | 30 (36%) | 157 (37%) |  |
| ≥30 (obese) | 22 (26%) | 434 (28%) |  | 22 (26%) | 110 (26%) |  |
| Not recorded | 17 (20%) | 419 (27%) |  | 17 (20%) | 87 (21%) |  |
| Comorbidities |  |  |  |  |  |  |
| Diabetes | 24 (29%) | 311 (20%) | 0.07 | 24 (29%) | 106 (25%) | 0.52 |
| Hypertension | 29 (35%) | 428 (28%) | 0.19 | 29 (35%) | 124 (30%) | 0.36 |
| CAD | 9 (11%) | 109 (7%) | 0.21 | 9 (11%) | 37 (9%) | 0.58 |
| Heart failure | 7 (8%) | 85 (6%) | 0.28 | 7 (8%) | 26 (6%) | 0.47 |
| ESRD or CKD | 11 (13%) | 130 (8%) | 0.14 | 11 (13%) | 47 (11%) | 0.62 |
| Chronic pulmonary disorders | 2 (2%) | 120 (8%) | 0.07 | 2 (2%) | 6 (11%) | 0.52 |
| Initial oxygen requirement |  |  | 0.39 |  |  | 0.85 |
| Room air | 25 (30%) | 378 (25%) |  | 25 (30%) | 116 (28%) |  |
| Nasal canula | 38 (45%) | 678 (44%) |  | 38 (45%) | 187 (44%) |  |
| Non-rebreather or similar | 21 (25%) | 480 (31%) |  | 21 (25%) | 117 (28%) |  |

BMI: body mass index; CAD: coronary artery disease; ESRD: end-stage renal disease; CKD: chronic kidney disease.

**Supplemental Table 2.** Final Cox proportional hazards model of risk factors for death or intubation among patients with COVID.

| Characteristics | Death or intubation /<br>n at risk (%) | Hazard ratio (95% CI) |  |
| --- | --- | --- | --- |
|  |  | Full model | Final model |
| Famotidine |  |  |  |
| No | 332 / 1,536 (22%) | Reference | Reference |
| Yes | 8 / 84 (10%) | 0.43 (0.21 – 0.86) | 0.42 (0.21-0.85) |
| Age |  |  |  |
| <50 years old | 19 / 333 (5.7%) | Reference | Reference |
| 50-65 years old | 75 / 514 (15%) | 2.94 (1.77-4.89) | 3.03 (1.83-5.03) |
| >65 years old | 246 / 773 (32%) | 7.51 (4.66-12.1) | 7.68 (4.79-12.3) |
| Sex |  |  |  |
| Male | 197 / 909 (22%) | Reference | --- |
| Female | 143 / 711 (20%) | 1.11 (0.89-1.38) |  |
| Race/ethnicity |  |  |  |
| Hispanic | 129 / 626 (21%) | Reference | --- |
| White, non-hispanic | 84 / 355 (24%) | 0.99 (0.75-1.31) |  |
| Black, non-hispanic | 59 / 340 (17%) | 0.82 (0.60-1.13) |  |
| Other | 68 / 299 (23%) | 1.14 (0.85-1.53) |  |
| Body mass index, kg/m<sup>2</sup> |  |  |  |
| <25.0 | 86 / 310 (28%) | Reference | --- |
| 25.0-29.9 (overweight) | 92 / 418 (22%) | 0.88 (0.65-1.18) |  |
| ≥30 (obese) | 89 / 456 (20%) | 0.97 (0.72-1.31) |  |
| Not recorded | 73 / 436 (17%) | 0.67 (0.49-0.92) |  |
| Comorbidities |  |  |  |
| Diabetes | 72 / 335 (21%) | 1.02 (0.75-1.37) | --- |
| Hypertension | 94 / 457 (21%) | 0.72 (0.54-0.97) | 0.74 (0.58-0.94) |
| CAD | 24 / 118 (20%) | 0.77 (0.49-1.21) | --- |
| Heart failure | 24 / 92 (26%) | 1.06 (0.67-1.67) | --- |
| ESRD or CKD | 33 / 141 (23%) | 1.16 (0.77-1.75) | --- |
| Chronic pulmonary disorders | 29 / 122 (24%) | 1.29 (0.87-1.93) | --- |
| Initial oxygen requirement |  |  |  |
| Room air | 52 / 403 (13%) | Reference | --- |
| Nasal canula | 155 / 716 (22%) | 1.60 (1.17-2.19) | 1.63 (1.19-2.24) |
| Non-rebreather | 133 / 501 (27%) | 2.48 (1.79-3.44) | 2.39 (1.73-3.29) |

CAD: coronary artery disease; ESRD: end-stage renal disease; CKD: chronic kidney disease.

